# Targeted vaccination and the speed of SARS-CoV-2 adaptation

**DOI:** 10.1101/2021.06.09.21258644

**Authors:** Sylvain Gandon, Sébastien Lion

## Abstract

The limited supply of vaccines against SARS-CoV-2 raises the question of targeted vaccination. Older and more sensitive hosts should be vaccinated first to minimize the disease burden. But what are the evolutionary consequences of targeted vaccination? We clarify the consequences of different vaccination strategies through the analysis of the speed of viral adaptation measured as the rate of change of the frequency of vaccine-escape mutations. We show that a vaccine-escape mutant is expected to spread faster if vaccination targets individuals which are likely to be involved in a higher number of contacts. We also discuss the pros and cons of dose-sparing strategies. Because delaying the second dose increases the proportion of the population vaccinated with a single dose, this strategy can both speed-up the spread of the vaccine-escape mutant and reduce the cumulated number of deaths. Hence, slowing down viral adaptation may not always be the optimal vaccination strategy. We contend that a careful assessment of the consequences of alternative vaccination strategies on both (i) the speed of adaptation and (ii) the mortality is required to determine which individuals should be vaccinated first.

---

The development of effective vaccines against SARS-CoV-2 raises hope regarding the possibility to eventually halt the ongoing pandemic. To cope with the limited supply of vaccines at a given time, different vaccination strategies have been proposed [1, 2, 3]. For instance, most countries prioritise older people, which are more at risk, or delay the second dose to maximise the number of partially vaccinated individuals. The cost-to-benefit ratio of these strategies depends on the goal we are trying to achieve. If we want to minimize disease and mortality we should target more sensitive and vulnerable hosts. If we want to minimize disease spread and epidemic size we may want to target hosts involved in more transmission. However, it is more difficult to assess the impact of these strategies on the evolution of vaccine-escape variants, which, worryingly, could erode the efficacy of vaccines.

Given that hosts differ both in their sensitivity to the disease and in their contribution to transmission, who should we vaccinate first if we want to minimise the risk of evolution of vaccine-escape mutants? Here, we try to answer this question using deterministic models that can accurately describe the dynamics of multiple variants after they successfully managed to reach a density at which they are less sensitive to the action of demographic stochasticity. We study the impact of different vaccination strategies on the rate of change of the frequency of vaccine-escape mutations, which allows us to quantify the speed of virus adaptation to vaccines.

## 1 Results

We are interested in tracking the frequency *p*_*m*_ of a vaccine-escape mutant in the viral population. It is possible to show that under a broad range of conditions one can approximate the dynamics of the vaccine-escape mutant frequency as:

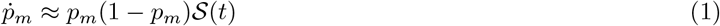

where 𝒮 (*t*) is the selection coefficient on the vaccine-escape mutation. This selection coefficient measures the rate of change of the logit of the frequency of the vaccine escape mutation and provides a relevant measure for the speed at which the viral population is adapting (see Methods).

In targeted vaccination strategies, we aim to preferentially vaccinate hosts with specific epidemiological characteristics. For instance, we could target hosts that have more contacts, or are more at risk of a severe disease. In our model, we therefore introduce some heterogeneity among hosts. As a result, from the point of view of the parasite, the *quality* of the host may differ among infected hosts, and this is likely to affect the dynamics of vaccine-escape mutations. To quantify host quality, we use the concept of reproductive value, a key concept in demography and evolutionary biology [4, 5, 6]. Reproductive value measures how much a virus infecting a given class of hosts will contribute to the future of the viral population. Our general mathematical analysis allows us to take the difference in host quality into account when calculating the selection coefficient *𝒮* (*t*) (see Methods). We use this approach to explore two different scenarios.

### Heterogeneity in contact numbers

In the first scenario we assume that hosts differ in their ability to mix and thus to transmit the disease. More specifically we assume that some hosts (*L*) have a low number of social interactions, while other hosts (*H*) have a higher number of contacts. This increased rate of social interactions is captured by a parameter *ℳ* ≥ 1. Susceptible hosts are initially naive (*S*^*L*^ and *S*^*H*^) but they can become vaccinated (*Ŝ* ^*L*^ and *Ŝ* ^*H*^) at rates *ν*^*L*^ and *ν*^*H*^, respectively. When vaccinated, hosts have reduced transmissibility and infectivity, but vaccine escape can erode these benefits. We consider different viral strains characterised by an escape trait *e* which takes values between 0 (no escape) and 1 (full escape). The capacity of a variant to reduce the effect of the vaccine on transmissibility (resp. infectivity) is captured by a function *E*_*τ*_ (*e*) (resp. *E*_*σ*_(*e*)), which allows us to quantify the overall ability of the virus to escape the protective effects of the vaccine as *E*(*e*) = *E*_*τ*_ (*e*)*E*_*σ*_(*e*).

In the Methods, we derive a simple approximation for the strength of selection acting on the vaccine-escape mutant:

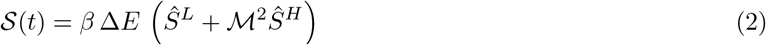

where Δ*E* refers to the change in vaccine escape ability caused by the mutation. This tells us that the intensity of selection depends on (i) the ability of the virus mutant to escape the protective effects of vaccine, (ii) the density of vaccinated hosts in the different types of hosts (*L* or *H*), and (iii) the relative number of contacts of each class of hosts. Note that the epidemiological impact of a higher contact rate (ℳ *>* 1) translates into a magnified selective impact (ℳ^2^). Thus, if we have to chose between vaccinating *L* and *H* hosts, targeting *H* hosts is expected to select more strongly for the vaccine-escape mutant. Figure 1A confirms that this approximation captures very well the temporal dynamics of the vaccine-escape mutant and can be used to understand the evolutionary consequences of different targeted vaccination strategies.

**Figure 1:**
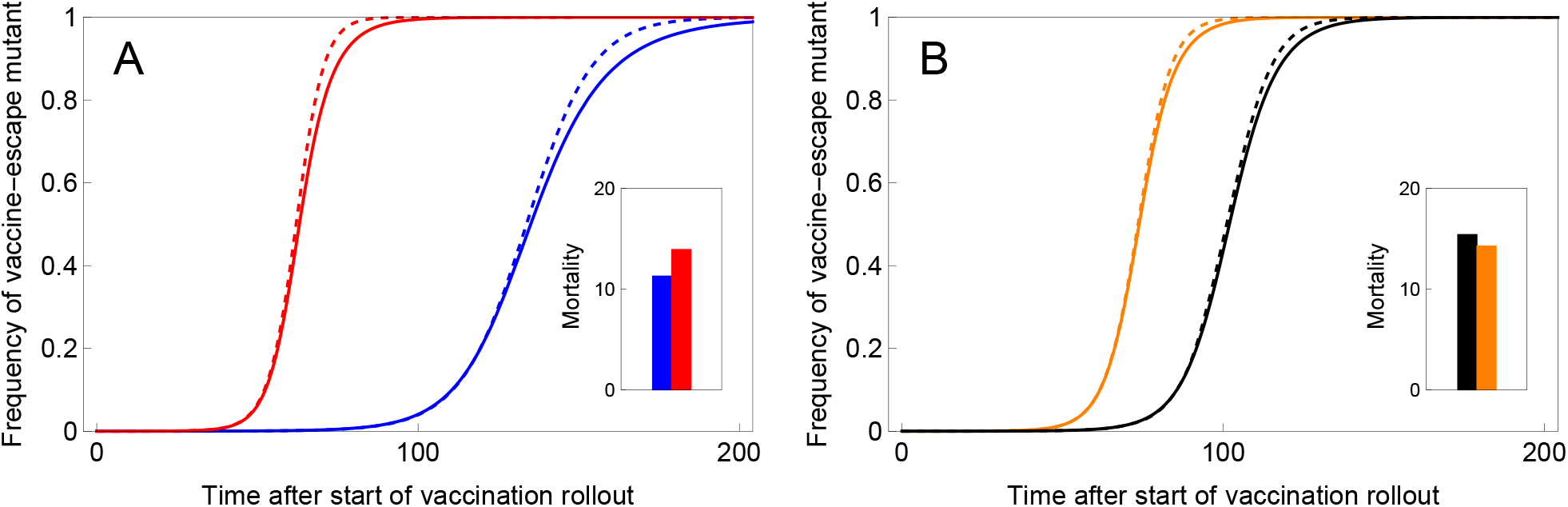
Targeted vaccination can alter the speed of viral adaptation. In (A) we plot the change in frequency of the vaccine-escape mutant after the start of vaccination when only the low transmitters – the *L* hosts – are vaccinated (blue curves) or only the high transmitters – the *H* hosts – are vaccinated (red curves). In (B) we plot the change in frequency of the vaccine-escape mutant after the start of vaccination when vaccinated hosts receive two doses sequentially (black curves) or when a single dose is used for each host (orange curve). The latter vaccination strategy yields imperfect immunity but leads to a larger density of vaccinated hosts. In both figures the dashed lines are obtained from the analytical approximation (equations (2) and (3) respectively) and the full lines are obtained from exact numerical simulations. The inset graph shows the cumulated number of deaths at time *t* = 200 under the different vaccination strategies (see Methods for parameter values).

Of course, the choice of the vaccination strategy should not be based solely on the reduction of the speed of adaptation to vaccines. Indeed, the best way to limit the spread of vaccine-escape mutations would be to adopt the worst epidemiological strategy: avoiding to use vaccines. Yet, we urgently need vaccines to save lives and halt the current pandemic. Tracking the cumulative number of deaths after the start of vaccination roll out allows us to identify the consequences of distinct targeted vaccination strategies on mortality (see Methods). Figure 1A shows that targeting *L* hosts is expected to decrease mortality because we assume that *L* hosts (e.g. older individuals with less contacts) are also associated with higher risks of dying from the infection. Hence, targeting *L* hosts makes sense both for epidemiological and evolutionary reasons.

### Heterogeneity in the number of vaccination doses

In our second scenario, we assume that the heterogeneity among hosts is determined by their vaccination status. We distinguish between unvaccinated hosts (*S*), hosts partially vaccinated with one dose (*Ŝ*^I^) and hosts fully vaccinated with two doses (*Ŝ*^II^). Using the same approach as before, we obtain the following expression for the selection coefficient:

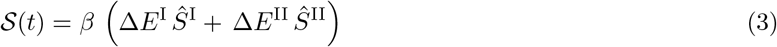

Equation (3) is very similar to Equation (2), but now we have to account for the fact that the escape mutation has different effects in each class. Hence, the influence of an increase in the densities of hosts vaccinated by a single or two doses of vaccines are weighted by Δ*E*^I^ and Δ*E*^II^, respectively. A single vaccine dose is likely to induce a lower protection against the virus (i.e. *E*^I^ *> E*^II^) but this does not necessarily imply that Δ*E*^I^ *>* Δ*E*^II^. In fact, we can show that if the vaccine is acting on a single step of the virus’ life cycle we expect Δ*E*^II^ *>* Δ*E*^I^. Delaying the acquisition of the second dose will have two effects: (i) a lower density *Ŝ*^II^ of fully vaccinated hosts decreases the more intense selection imposed by these hosts, (ii) but delaying the second dose allows for more hosts to be vaccinated and to increase *Ŝ*^I^ and may thus increase selection for the vaccine-escape mutant. We show in Figure 1B that this second effect can be more important than the first one and delaying the second dose can result in faster adaptation. However, the mortality may be lower because a larger fraction of the population would benefit from the protection of the vaccine. Hence, in contrast to the previous scenario, the strategy that maximises the speed of adaptation may result in a lower mortality. A more comprehensive exploration of the robustness of these conclusions is needed before making specific recommendations on delaying the second dose. Yet, the contrast between our two scenarios illustrates the necessity to quantify both the epidemiological and the evolutionary consequences of different targeted vaccination strategies to identify the optimal way to distribute vaccines.

## 2 Discussion

The speed of the spread of SARS-CoV-2 variants has baffled the scientific community [7, 8]. In spite of a relatively small mutation rate [9, 10] SARS-CoV-2 has the ability to produce mutations with variable phenotypic effects that fuel the adaptation to human populations. The growing concern regarding the ability of the virus to escape host immunity calls for tools that may allow us to anticipate the speed of the spread of vaccine-escape mutants. We show here that heterogeneity in the behaviour (scenario 1) and/or immune status (scenario 2) can induce variation in the strength of selection for vaccine-escape mutations. We contend that it is important to quantify this variation because it could be used to carry out targeted vaccination strategies that, for a given vaccination coverage, could limit the speed of adaptation of the virus.

We show that targeted vaccination on older hosts which are associated with lower number of contacts but higher risks of mortality is a good strategy to reduce both the spread of the vaccine-escape variant and the cumulative number of death. Gog et al. [3] used a different approach to identify vaccination strategies that could reduce what they call ‘vaccine escape pressure’, a quantity proportional to the number of infectious cases in vaccinated hosts. In contrast to our study, they conclude that vaccinating most of the vulnerable hosts and few of the mixers could be the most risky for vaccine escape. Yet, the criteria used in [3] is meant to capture the within-host selection pressure to escape immunity in vaccinated hosts. The link between this criteria and the change in frequency of the vaccine-escape mutant in the host population remains to be explored.

We also discuss the effect of delaying the second dose of the vaccine on viral adaptation and on mortality. In a recent model, Saad-Roy et al. [2] found that imperfect immunity induced by a single dose may lead to stronger within-host selection for vaccine-escape variants. In contrast, we focus on between-host selection and ask whether vaccine-escape variants can increase in frequency at the population level. Indeed, within-host selection is likely to constrain the emergence of vaccine-escape mutations in the first place but we contend that once those vaccine-escape mutants reach a significant fraction of the population, the fate of those mutations will be driven by between-host selection. Our analysis shows that the speed of viral adaptation critically depends on the balance between the within-host effects (i.e. the relative magnitudes of Δ*E*^I^ and Δ*E*^II^) and the population-level effects (i.e. the relative densities of hosts with one or two doses of the vaccine). We show that a higher speed of adaptation may be the price to pay for a reduced number of deaths (Figure 1B). Indeed, delaying the second dose allows us to protect (albeit partially) a larger fraction of the population (see [1] for an exploration of this effect). This positive effect can outweigh the negative consequences of an erosion of vaccine efficacy due to viral adaptation.

Vaccination is urgently needed to control the SARS-CoV-2 pandemic but viral adaptation could erode the efficacy of vaccines. Targeted vaccination may provide a way to delay this adaptation but, as illustrated with the second scenario, a strategy that minimizes the cumulated number of deaths may not necessarily minimize the speed of adaptation. Hence, as for any therapeutic interventions that may result in the evolution of pathogen resistance, the identification of an optimal vaccination strategy that reduces the death toll of the pandemic requires specific models accounting for both the epidemiology and the evolution of the virus.

## Data Availability

There is no data for this manuscript.

## Methods

### General approach

We first give a general overview of the method used to calculate the selection coefficient in structured host populations. The dynamics of hosts infected by pathogen strain *i* can be captured by a matrix **R**_*i*_ collecting the transition rates between host classes. Assuming that mutations have small phenotypic effects (i.e. *e*_*m*_ = *e*_*w*_ + *ε*), we can write the change in frequency of the mutant strain as

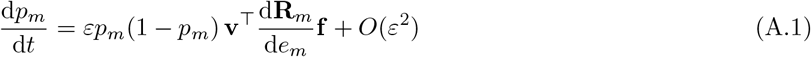

where **v**^⊤^is the vector of reproductive values and **f** is the vector of class frequencies. These vectors are conormalised such that **v**^⊤^**f** = 1 and satisfy the following dynamical equations

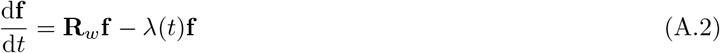

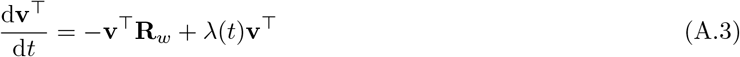

where **R**_*w*_ is the transition matrix for the wild-type strain and *λ*(*t*) is the per-capita growth rate of the resident population at time *t* (see [6, 11] for a more detailed description). For small *ε*, the mutant frequency *p*_*m*_ changes slowly compared to the ecological variables **f** and **v**, and we can use a quasi-equilibrium approximations obtained by setting the right-hand-sides of equations (A.2) and (A.3) to zero. This allows us to obtain analytical expressions for the class frequencies and reproductive values and thus to calculate the selection coefficient for a specific life cycle (scenario 1 vs scenario 2). Note that, although the weak selection assumption (small *ε*) is driving the separation of time scale, the approximation remains good when selection is strong as discussed in the two scenarios below.

### Scenario 1: should we preferentially vaccinate individuals with more contacts?

We assume that susceptible hosts in class *k* are produced at rate *θ*^*k*^ and die at rate *d*^*k*^ (where *k* = *L* or *H*). New susceptible hosts are vaccinated with probability *ν*^*k*^ representing the intensity of the vaccination campaign for each host class. We note ℳ *>* 1 the relative number of contacts of *H* hosts compared to *L* hosts and *ρ*_*τ*_ (resp. *ρ*_*σ*_) the relative transmissibility (resp. susceptibility) of vaccinated hosts compared to naive hosts of the same class. Both *ρ*_*τ*_ and *ρ*_*σ*_ are functions of the vaccine escape trait. With these assumptions, the force of infection of a pathogen strain *i* due to naive infected hosts is 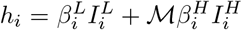, and 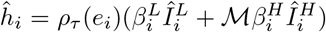 for vaccinated infected hosts. Note that vaccinated hosts are indicated by a “hat” (denoting protection). Hosts in class *k* infected by pathogen strain *i* eventually leaves the class at rate 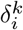 (resp. 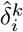 for vaccinated hosts) and can either recover or die. We assume that the probability 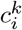 (resp 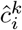) of dying after leaving the class 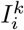 (resp. 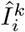) may depend on the host class *k* and pathogen strain *i* and we track the cumulated number of deaths *D*. This quantity can be used to compare the efficacy of different vaccination strategies. Note however that the probabilities 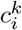 and 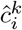 have no impact on evolutionary dynamics because these events occur when the host is assumed to be no longer infectious and consequently they do not affect pathogen fitness.

This yields the following dynamical system (see also figure A.1):

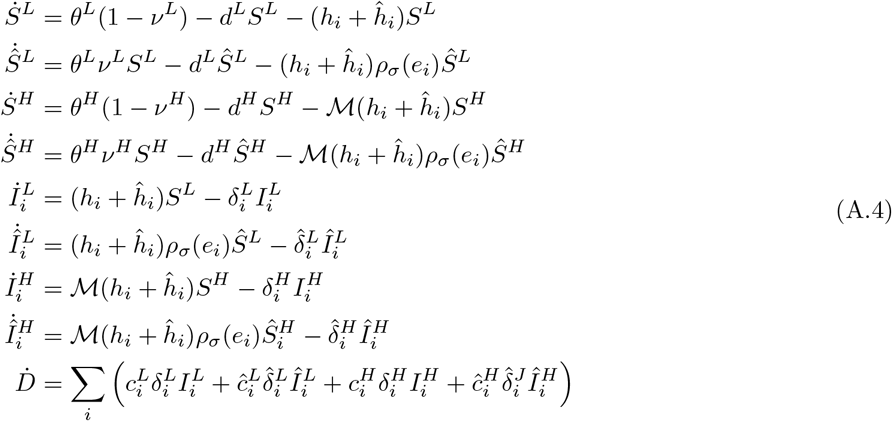

We analyse this general model under two simplifying but reasonable assumptions.

i. We assume that the pathogen strains only differ through their effect on the parameters *ρ*_*τ*_ and *ρ*_*σ*_ (that is, we only look at vaccine escape mutations, not mutations that can also affect transmissibility or virulence).
ii. We assume that host classes *L* and *H* only differ through their number of contacts, so that *β*^*L*^ = *β*^*H*^ = *β* (same baseline transmissibility) and 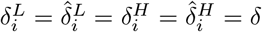.

The latter assumption implies that the duration of infection is the same in all classes, but the effect of vaccination and host behaviour can be captured through the probabilities *c*^*L*^, *ĉ*^*L*^, *c*^*H*^, *ĉ*^*H*^ (again, note that there is no influence of the pathogen genotype on disease outcome). For instance, in our simulations, we assume that *L* hosts tend to have less contacts but a higher mortality risk, while *H* hosts have more contacts but a lower mortality risk. This may reflect the observed differences between age

The transition matrix **R**_*i*_ is the 4 × 4 matrix of per-capita transition rates of the pathogen between the 4 different types of hosts:

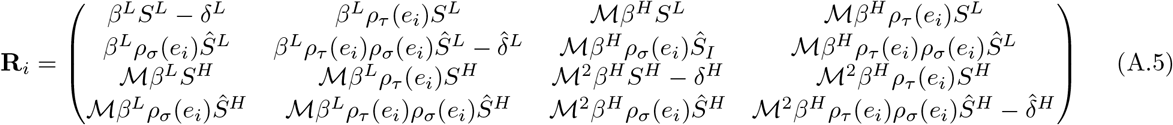

We are interested in the dynamics of the frequency of the vaccine-escape mutant, which is:

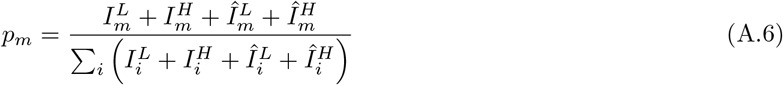

The dynamics of *p*_*m*_ can be calculated by plugging the expressions of **R**_*w*_ and **R**_*m*_ into equations (A.1), (A.2) and (A.3). We obtain, after some rearrangements

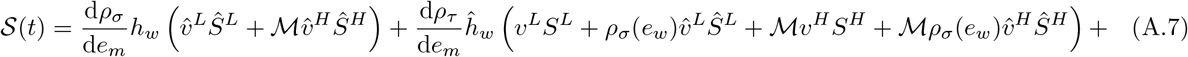

where the vector 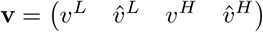 collects the reproductive values of an individual resident pathogen in classes *I*^*L*^, *Î*^*L*^, *I*^*H*^ and *Î*^*H*^ respectively. Note that this only result only depends on assumption (i) above.

It is possible to simplify the expression of the selection coefficient by treating the reproductive values as fast variables. In particular, using our assumption (ii), this leads to the following quasi-equilibrium approximations:

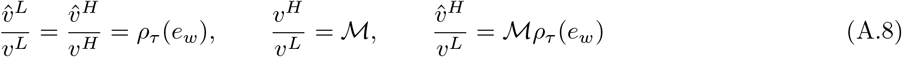

Similarly, we have the following quasi-equilibrium approximations for the class frequencies, which give the fraction of infected individuals in a given class:

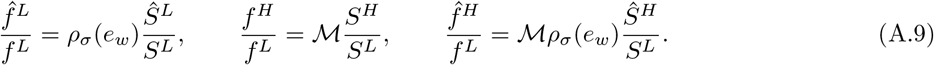

Together with the normalisation condition 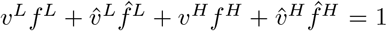, we can use these relationships to obtain:

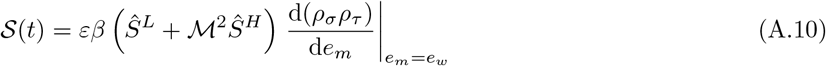

To recover Equation (2) in the main text, we use the notations *E*_*τ*_ = *ρ*_*τ*_, *E*_*σ*_ = *ρ*_*σ*_, *E* = *E*_*τ*_ *E*_*σ*_ and

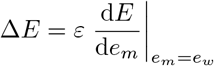

which is the first order approximation of the difference *E*(*e*_*m*_) − *E*(*e*_*w*_).

#### Numerical simulations

In our applications, we use a linear model of vaccine escape:

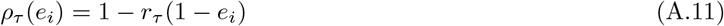

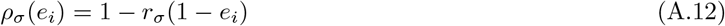

where *r*_*τ*_ and *r*_*σ*_ give the vaccine efficacy in the absence of vaccine escape mutation (i.e. *e*_*i*_ = 0). When *e*_*i*_ = 1 (full vaccine escape), the vaccine offers no reduction in transmissibility and susceptibility (*ρ*_*σ*_ = *ρ*_*τ*_ = 1). We assume that vaccination can also protect against disease induced mortality and we define *r*_*µ*_ so that *ĉ*^*L*^ = (1 − *r*_*µ*_)*c*^*L*^ and *ĉ*^*H*^ = (1 − *r*_*µ*_)*c*^*H*^.

Initial conditions used in Figure 1A in the main text: *S*^*L*^(0) = *S*^*H*^ (0) = 1, *Ŝ*^*L*^ = *Ŝ*^*H*^ = 0, *D*(0) = 0, *I*^*L*^(0) = *I*^*H*^ (0) = 0.1, *Î*^*L*^(0) = *Î*^*H*^ (0) = 10^−7^, *p*_*m*_(0) = 10^−4^. Vaccination starts at *t* = 1000 and the parameters used in Figure 1A in the main text are the following:

| Parameter | Value |  |
| --- | --- | --- |
| $\theta^L = \theta^H$ | 2 | |
| $d^L = d^H$ | 0.01 | |
| $\delta$ | 1 | |
| $\beta$ | 0.01 | |
| $c^L$ | 0.02 | |
| $c^H$ | 0.001 | |
| $r_\sigma$ | 0.8 | |
| $r_\tau$ | 0 | |
| $r_\mu$ | 0.9 | |
| $\mathcal{M}$ | 2 | |
| $e_w$ | 0 | |
| $e_m$ | 0.5 | |
| $\nu^L$ | 0.25 (blue) | 0.005 (red) |
| $\nu^H$ | 0.005 (blue) | 0.25 (red) |

**Figure A.1:**
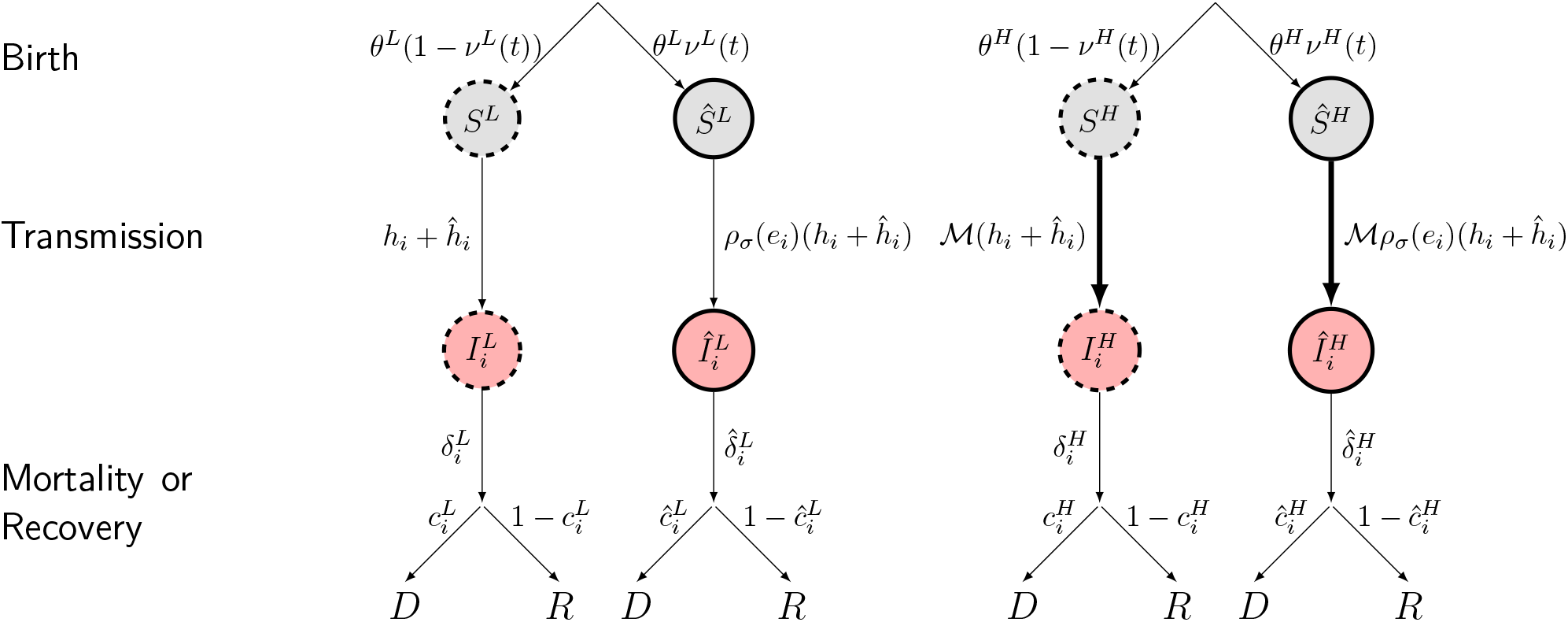
A schematic representation of the model. New uninfected hosts are produced at a rate *λ*^*k*^ and are vaccinated with probability *ν*^*k*^, yielding uninfected hosts (*S*^*k*^ and *Ŝ*^*k*^) which die at a rate *d* while infected hosts (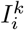 and 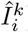) die or recover at rates 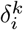 and 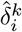, where *i* refers to the virus genotype: the wild type (*i* = *w*) or the escape mutant (*i* = *m*). The rate of infection of naive hosts by the genotype *i* depends on the forces of infection due to naive (*h*_*i*_) or vaccinated hosts 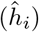 as detailed in the text. Vaccination reduces the force of infection and *e*_*i*_ refers to the ability of the genotype *i* to escape the immunity triggered by vaccination (we assume *e*_*m*_ *> e*_*w*_). After infection, hosts can either die or recover, with probabilities 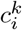 and 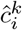.

### Scenario 2: should we delay the second dose?

For our second scenario, we consider three classes of susceptible hosts: unvaccinated (*S*), vaccinated with 1 dose (*S*^I^) and vaccinated with 2 doses (*S*^II^). Unvaccinated susceptible hosts are produced at rate *θ* and can be given a first dose of vaccine at rate *ν*^I^. Susceptible hosts that have received one dose can be given a second dose at rate *ν*^II^. With one dose, the relative transmissibility (resp. susceptibility) of vaccinated hosts with respect to pathogen strain *e*_*i*_ is 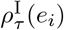 (resp. 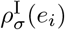). With two doses, we use the notation 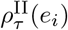 and 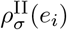. Apart from these assumptions, the life cycle is similar to the one used for scenario 1, and we have the following dynamics (see also figure A.2):

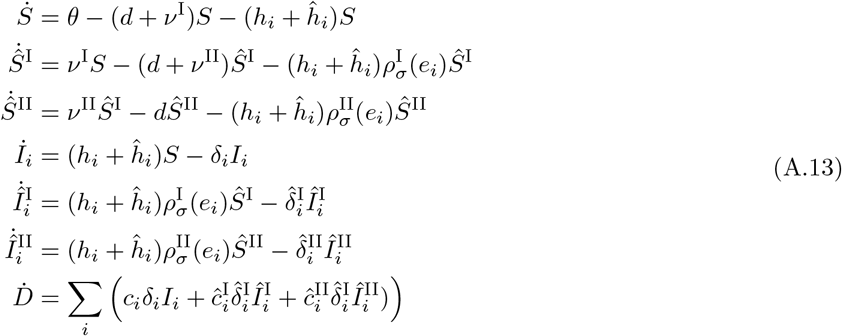

where the forces of infection by virus strain *i* are *h*_*i*_ = *β*_*i*_*I*_*i*_ and 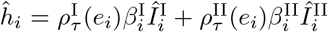. For simplicity, we will also assume, as in scenario 1, that 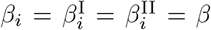 and 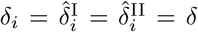, so that (1) hosts only differ through the parameters *ρ*_*τ*_ and *ρ*_*σ*_, and (2) the viral strains only differ through the parameters *ρ*_*τ*_ and *ρ*_*σ*_. We also assume that 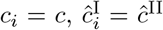 and 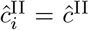 to account for potential differences between mortality rates between different classes of hosts (but no influence of the pathogen genotype).

With these assumptions, the matrix **R**_*i*_ is

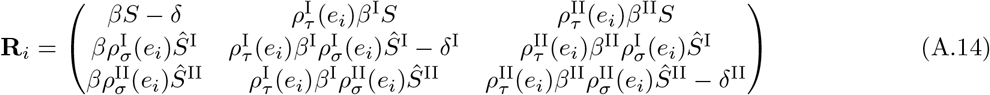

and the quasi-equilibrium relationships for class frequencies and reproductive values, when *β*^I^ = *β*^II^ = *β* and 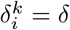 are:

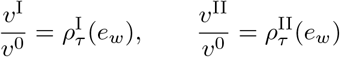

and

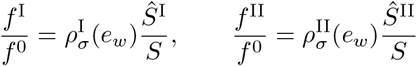

where **v** = *v*^0^ *v*^I^ *v*^II^ and **f** = *f* ^0^ *f* ^I^ *f* ^II^. Together with the normalisation condition *v*^0^*f* ^0^ + *v*^I^*f* ^I^ + *v*^II^*f* ^II^ = 1, these relationships allow us to rearrange equation (A.1) to obtain

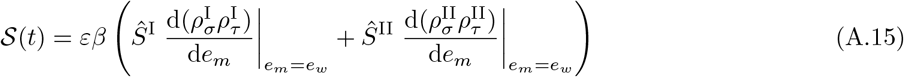

which is Equation (3) in the main text using the same notations as in Scenario 1.

#### Numerical simulations

We use the same linear model of vaccine escape as in scenario 1, but we allow for different vaccine efficacies depending on the number of doses:

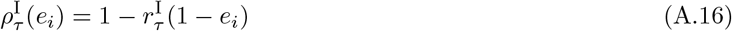

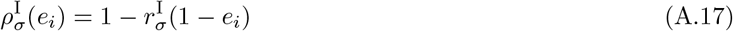

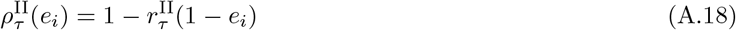

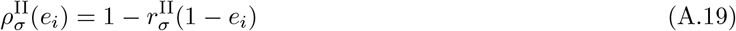

where *r*_*τ*_ and *r*_*σ*_ give the vaccine efficacy in the absence of vaccine escape mutation (i.e. *e*_*i*_ = 0). When *e*_*i*_ = 1 (full vaccine escape), the vaccine offers no reduction in transmissibility and susceptibility (*ρ*_*σ*_ = *ρ*_*τ*_ = 1). We assume that vaccination can also protect against disease induced mortality and we define *r*_*µ*_ so that 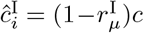 and 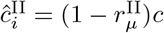.

Initial conditions in Figure 1B in the main text: *S*(0) = *Ŝ*^I^(0) = *Ŝ*^II^(0) = 1, *D*(0) = 0, *I*(0) = 0.1, *Î*^I^(0) = *Î*^II^(0) = 10^−7^, *p*_*m*_(0) = 10^−4^. Vaccination starts at *t* = 1000 and the parameters used in Figure 1B in the main text are the following:

| Parameter | Value |  |
| --- | --- | --- |
| $\theta$ | 4 | |
| $d$ | 0.01 | |
| $\delta$ | 1 | |
| $\beta$ | 0.01 | |
| $c$ | 0.01 | |
| $r_\sigma^I$ | 0.6 | |
| $r_\sigma^{II}$ | 0.9 | |
| $r_\tau^I = r_\tau^{II}$ | 0 | |
| $r_\mu^I$ | 0.5 | |
| $r_\mu^{II}$ | 0.9 | |
| $e_w$ | 0 | |
| $e_m$ | 0.5 | |
| $\nu^I$ | 0.01 (black) | 0.02 (orange) |
| $\nu^{II}$ | 0.01 (black) | 0.00001 (orange) |

**Figure A.2:**
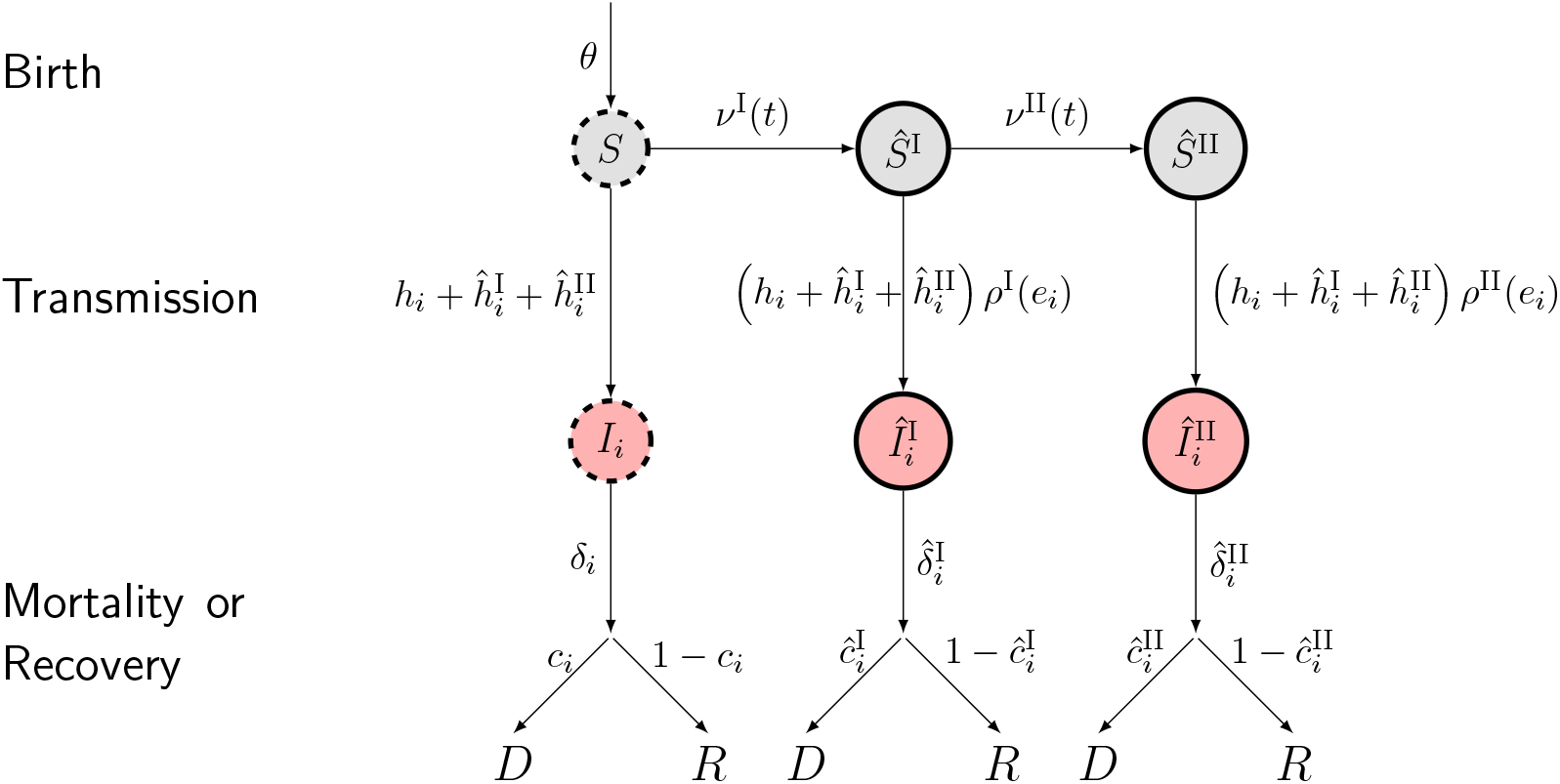
A schematic representation of the second scenario. Naive and uninfected hosts (*S* hosts) are introduced at a rate *θ* and are vaccinated at rate *ν*^I^ later on. Hosts vaccinated with one dose (*Ŝ*^I^) receive a second dose at rate *ν*^II^. Uninfected hosts (*S, Ŝ*^I^ and *Ŝ*^II^) die at a rate *d* (unshown) while infected hosts (*I*_*i*_, 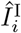 and 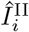) die at rates *δ*_*i*_, 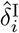 and 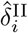 respectively, where *i* refers to the virus genotype: the wild type (*i* = *w*) or the escape mutant (*i* = *m*). The rate of infection of naive hosts by the genotype *i* depends on the forces of infection *h*_*i*_, 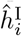 and 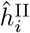 (see text for details). Vaccination reduces the force of infection and *e*_*i*_ refers to the ability of the genotype *i* to escape the immunity triggered by vaccination (we assume *e*_*m*_ *> e*_*w*_). After infection, hosts can either die or recover, with probabilities *c*_*i*_, 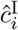 and 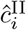.

